# Attention Is All You Need: A Universal Augmented Reality Surgical Navigation Framework

**DOI:** 10.64898/2026.09.20.26363284

**Authors:** Danyang Zhao, Yu Qiu, Yue Guo, Xutao Luo, Yuhan Liu, Xinru Li, Hui Chen, Xiaoxiao Cai, Taoran Tian

## Abstract

Hard-tissue surgery requires precise spatial control, while conventional computer-assisted navigation presents guidance separately from the operative field. Augmented reality can bring navigation information closer to the surgeon, but how best to translate this information into intuitive intraoperative guidance remains underexplored. We therefore developed an augmented reality navigation system (ANS) that translates real-time deviations from the planned trajectory into co-located holographic action cues. Evaluation comprised typodont and ex vivo porcine experiments, a randomized crossover study in 40 dental trainees, and eight clinical cases. In porcine mandibles, ANS showed lower angular deviation than static and dynamic navigation (1.78° vs 6.42° and 4.02°). Among trainees, accuracy was comparable with dynamic navigation, while workload was lower (29.18 vs 33.15), usability higher (73.13 vs 68.88), and visual-focus ratings improved (3.55 vs 2.28). Clinical cases supported feasibility. These findings support co-located action guidance as an alternative interface paradigm for surgical navigation.

## Introduction

Hard-tissue surgery requires submillimeter precision, yet limited visibility and complex anatomy can produce mean drilling errors of 1.3-6.33 mm during surgical procedures ^1–3^. Deviations of this magnitude may injure critical anatomical or neurovascular structures and compromise patient safety. Computer-assisted surgical navigation (CAS) has therefore become an important strategy for reducing intraoperative variability in head-and-neck ^4–6^, spinal ^7–9^ and neurosurgical procedures ^10,11^.

Despite these gains, conventional CAS may still disrupt the surgeon’s natural hand-eye coordination. Navigation information is usually displayed away from the operative field, requiring repeated gaze shifts between the patient and an external monitor. Surgeons must also translate multiplanar images, coordinates and trajectory lines into concrete manual actions. This separation between information and action increases visual switching, cognitive mapping demands and procedural risk during critical steps ^12,13^.

Augmented reality (AR) may address part of this problem by placing digital navigation information within the surgeon’s field of view. AR guidance has been explored for pedicle screw placement ^14,15^, fracture fixation ^16,17^ and total shoulder arthroplasty ^18^. In pedicle screw insertion, for example, AR-guided navigation has reported accuracy rates of 94.1-97.4%, compared with 89.3% for freehand insertion and 92.3% for robotic-assisted navigation ^19^. A recent study in *npj Digital Medicine* suggests that head-mounted AR navigation can achieve accuracy comparable to conventional navigation while improving user experience, although limitations in visualization and clinical generalizability remain ^20^.

Most medical AR systems follow two paradigms (**Figure 1a**). The virtual-monitor paradigm projects floating panels near the surgical field to display patient data, navigation cues and anatomical models^21–25^. The three-dimensional overlay paradigm uses a head-mounted display to align virtual anatomy or implant positions with the operative site ^26,27^. Both paradigms reduce some attention diversion, but neither fully removes the need for cognitive mapping. Virtual monitors still require symbolic interpretation, whereas three-dimensional overlays can introduce clinically relevant depth and distance errors through vergence-accommodation conflict ^28^. A practical AR paradigm that converts navigation data directly into action-level guidance is therefore still needed.

**Figure 1.**
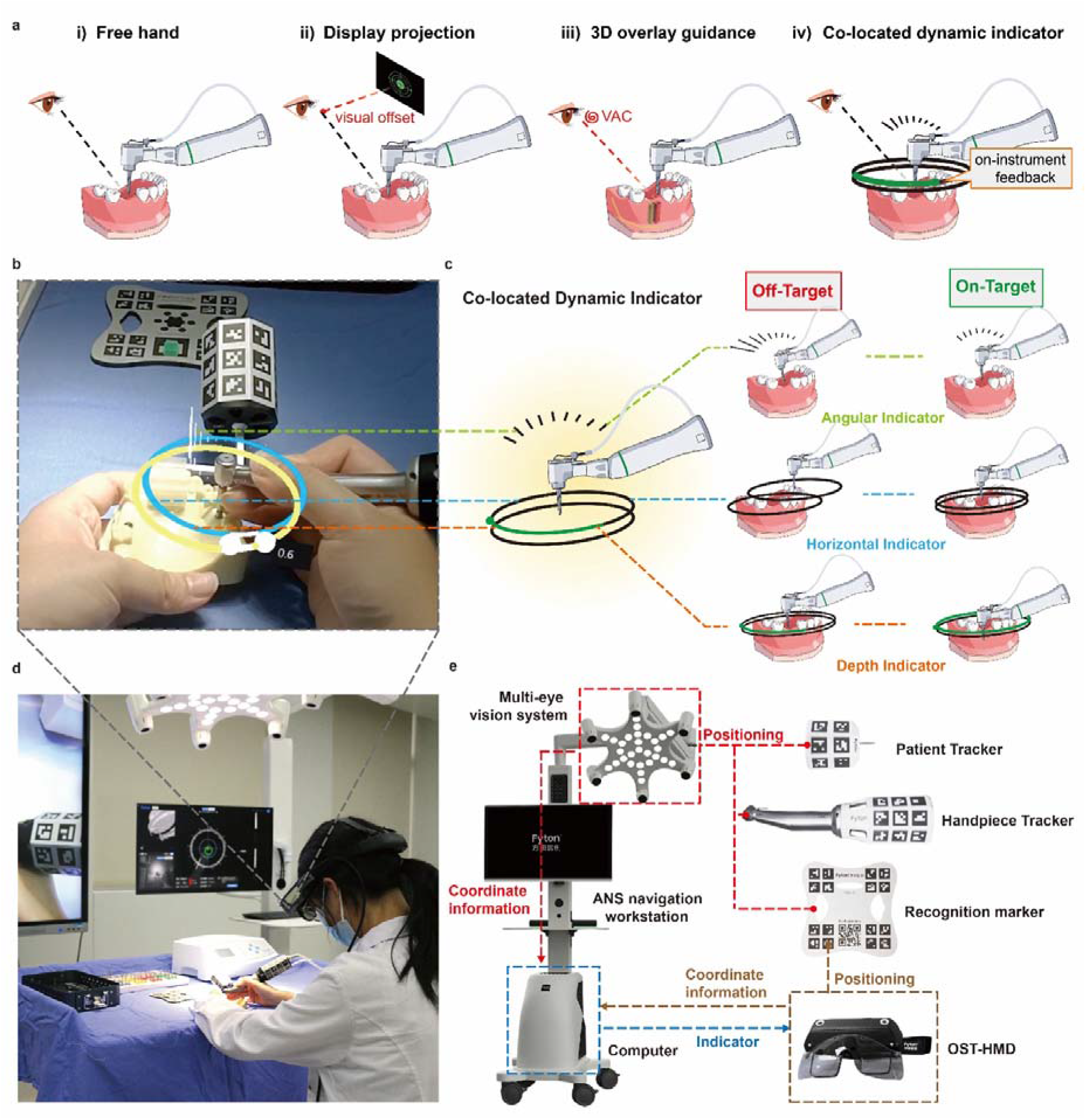
Concept, Visual Guidance, and Hardware Architecture of the Augmented Reality–Based Navigation System. (a) Comparison of freehand surgery and 3 augmented reality guidance paradigms. Display-projection systems present navigation information on a virtual screen and still require visual switching and interpretation. Three-dimensional overlay systems align virtual anatomical information with the operative site but may be affected by vergence-accommodation conflict. The co-located dynamic indicator converts navigation errors into action-oriented cues positioned at the surgical instrument and operative field. (b) First-person view of the holographic indicators during ANS-guided implant osteotomy preparation in a typodont model. (c) Operating principles of the co-located dynamic indicator. Angular deviation is represented by the direction and length of the radial indicator, horizontal deviation by the relative positions of the handpiece-anchored and site-anchored rings, and drilling depth by the movement of the depth indicator. Alignment of the visual elements indicates an on-target instrument position and trajectory. (d) Operating-room setup for ANS-guided implant placement. (e) Hardware and information-flow architecture of the ANS, including the multi-eye vision system, navigation workstation, computer, patient tracker, handpiece tracker, recognition marker, and optical see-through head-mounted display. Positioning and coordinate information are processed by the navigation system and transmitted to the head-mounted display for real-time indicator rendering.

We propose a dynamic holographic indicator that is co-located with the surgical instrument. This design converts navigation data into directly perceptible spatial cues, reducing the cognitive work needed to map abstract information onto manual actions ^29–31^. Because the indicator remains anchored to the operative field, it also limits gaze shifts and avoids the visual competition caused by external displays or occluding virtual models ^32,33^. The aim is not only to present more information, but to shorten the pathway from perception to execution.

On this basis, we developed an augmented-reality navigation system (ANS) and evaluated it in dental implant placement, a representative hard-tissue puncture procedure that requires simultaneous control of implant position, angulation and depth. We compared ANS with freehand surgery (FHS), static computer-assisted implant surgery (s-CAIS) and dynamic computer-assisted implant surgery (d-CAIS) in typodont models, ex vivo porcine mandibles and a preliminary clinical first-molar case series in humans.

We assessed placement accuracy, procedural efficiency and user experience to determine whether co-located action guidance can support precise navigation while reducing cognitive burden.

## Results

### Development of a co-located augmented reality navigation system

We developed a co-located augmented reality navigation system (ANS) that integrates real-time optical tracking with holographic action guidance directly within the surgeon’s field of view (Figure 1). Unlike virtual-monitor or three-dimensional overlay paradigms, ANS translates deviations from the planned trajectory into instrument-centred visual cues that indicate the corrective action required during drilling (**Figure 1a, b**).

The holographic indicator provides coordinated guidance for lateral position, angulation and drilling depth. Angular deviation is represented by the direction and extent of the radial indicator, lateral deviation by the relative alignment of the handpiece-anchored and site-anchored visual elements, and remaining drilling depth by a depth indicator. Alignment of these elements indicates alignment of the instrument with the planned position and trajectory (**Figure 1c**).

The resulting system integrates a self-developed optical see-through head-mounted display, distributed visible-light tracking, marker-based tracking of the patient and surgical handpiece, and real-time visualization of the surgical model and instrument pose (**Figure 1d, e**). Together, these components allow navigation information to remain co-located with the operative field throughout the drilling procedure.

### ANS improves hard-tissue puncture accuracy in typodont models

ANS produced significantly lower coronal, apical and angular deviations than FHS (**Figure 2h-r**, **Table 1**). Angular deviation decreased from 5.12 degrees with FHS to 2.03 degrees with ANS (P < 0.001), and apical deviation decreased from 1.67 mm to 0.79 mm (P < 0.001). Coronal deviation was 0.81 mm with ANS versus 1.34 mm with FHS (P < 0.001).

**Figure 2.**
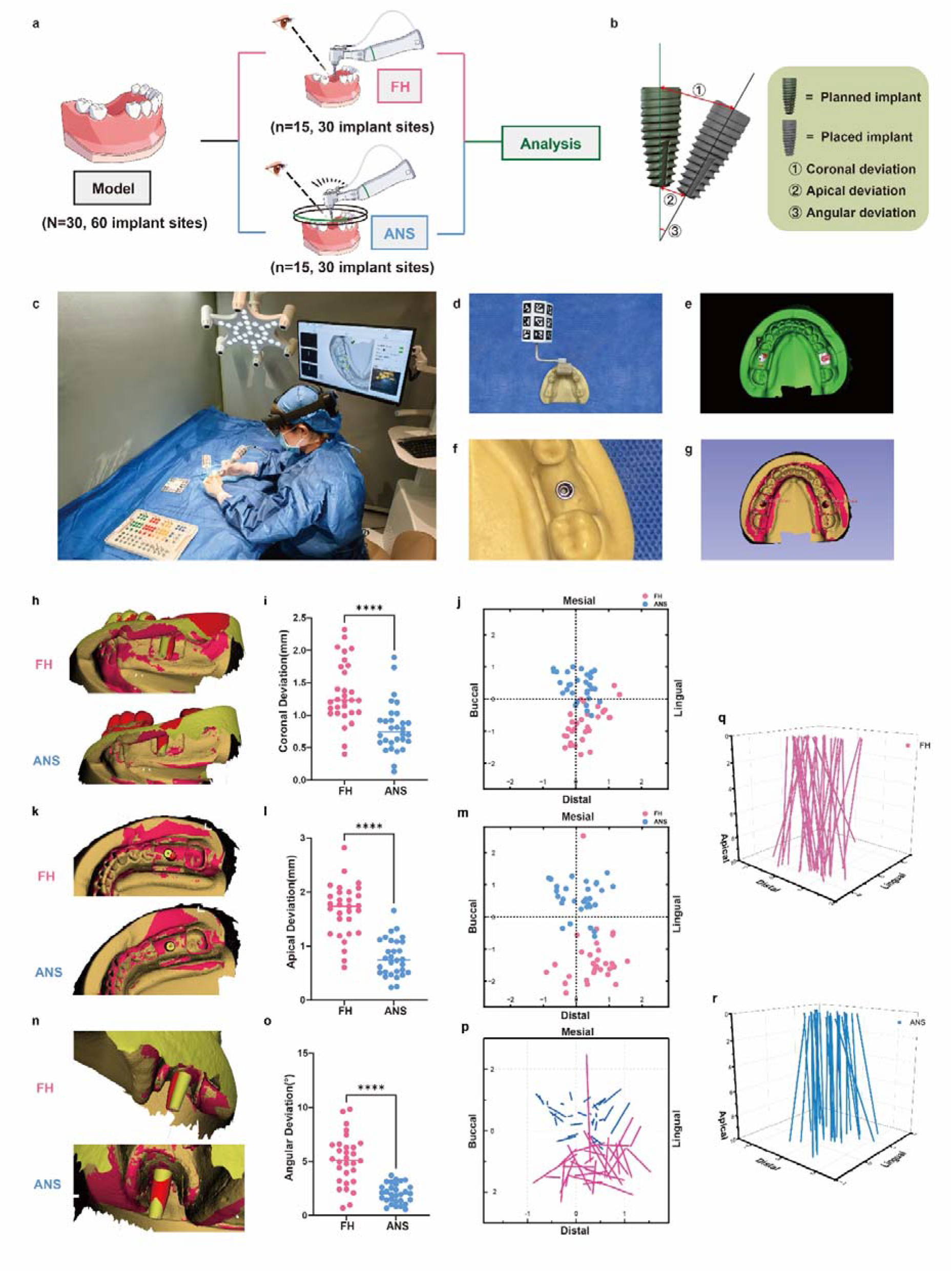
Accuracy evaluation of ANS-guided implant osteotomy in a dental model. (a) Schematic illustration of the experimental design. Implant osteotomies were performed in dental models using either freehand surgery (FH) or the augmented reality-based navigation system (ANS), followed by postoperative accuracy analysis. (b) Definition of coronal, apical, and angular deviations between the planned and placed implant trajectories. (c) Representative setup of ANS-guided drilling in the model experiment. (d–g) Workflow for accuracy assessment, including model tracking, preoperative planning, osteotomy preparation, and postoperative registration of planned and actual trajectories. (h–j) Representative reconstructed images, quantitative comparison, and directional distribution of coronal deviation in the FH and ANS groups. (k–m) Representative reconstructed images, quantitative comparison, and directional distribution of apical deviation. (n–p) Representative reconstructed images, quantitative comparison, and directional distribution of angular deviation. (q, r) Three-dimensional distribution of drilling trajectories in the FH and ANS groups. Data are presented as individual values with mean lines. Statistical significance was determined using unpaired t-tests or Mann–Whitney U tests according to data distribution. ****P < 0.0001.

**Table 1.** Deviation statistics for indicator-assisted ANS and FHS in typodont implant models. Values summarise angular, three-dimensional entry-point and three-dimensional apical deviations;

| Group | ANS<br>(n=30) | FHS<br>(n=30) | P value<br>(independent t-test) |
| --- | --- | --- | --- |
| **Angular deviation (degree)** |  |  |  |
| Mean±SD | 2.03±0.89 | 5.12±2.30 | <0.001 |
| Median | 1.99 | 5.13 |  |
| Min-Max | 0.57-3.71 | 0.70-9.83 |  |
| **3D Entry point deviation (mm)** |  |  |  |
| Mean±SD | 0.81±0.39 | 1.34±0.48 | <0.001 |
| Median | 0.75 | 1.23 |  |
| Min-Max | 0.13-1.89 | 0.40-2.32 |  |
| **3D Apical deviation (mm)** |  |  |  |
| Mean±SD | 0.79±0.35 | 1.67±0.49 | <0.001 |
| Median | 0.75 | 1.74 |  |
| Min-Max | 0.24-1.66 | 0.61-2.82 |  |
| Independent t-test was performed at P value<0.05; ANS, augmented reality-based navigation system; FHS, freehand surgery; SD, standard deviation; Min, minimum value; Max, maximum value. |  |  |  |

### ANS maintains accuracy in preclinical ex vivo testing

In ex vivo porcine mandibles, ANS showed lower angular deviation than s-CAIS and d-CAIS (1.78 ± 0.79 degrees vs. 6.42 ± 3.01 degrees and 4.02 ± 2.16 degrees, P < 0.001). ANS also showed lower apical deviation (0.82 ± 0.35 mm vs. 1.67 ± 0.77 mm and 1.19 ± 0.46 mm, P = 0.003). Entry-point deviation did not differ significantly, although ANS had the smallest mean value (0.79 ± 0.34 mm; **Figure 3h-r**, **Table 2**).

**Figure 3.**
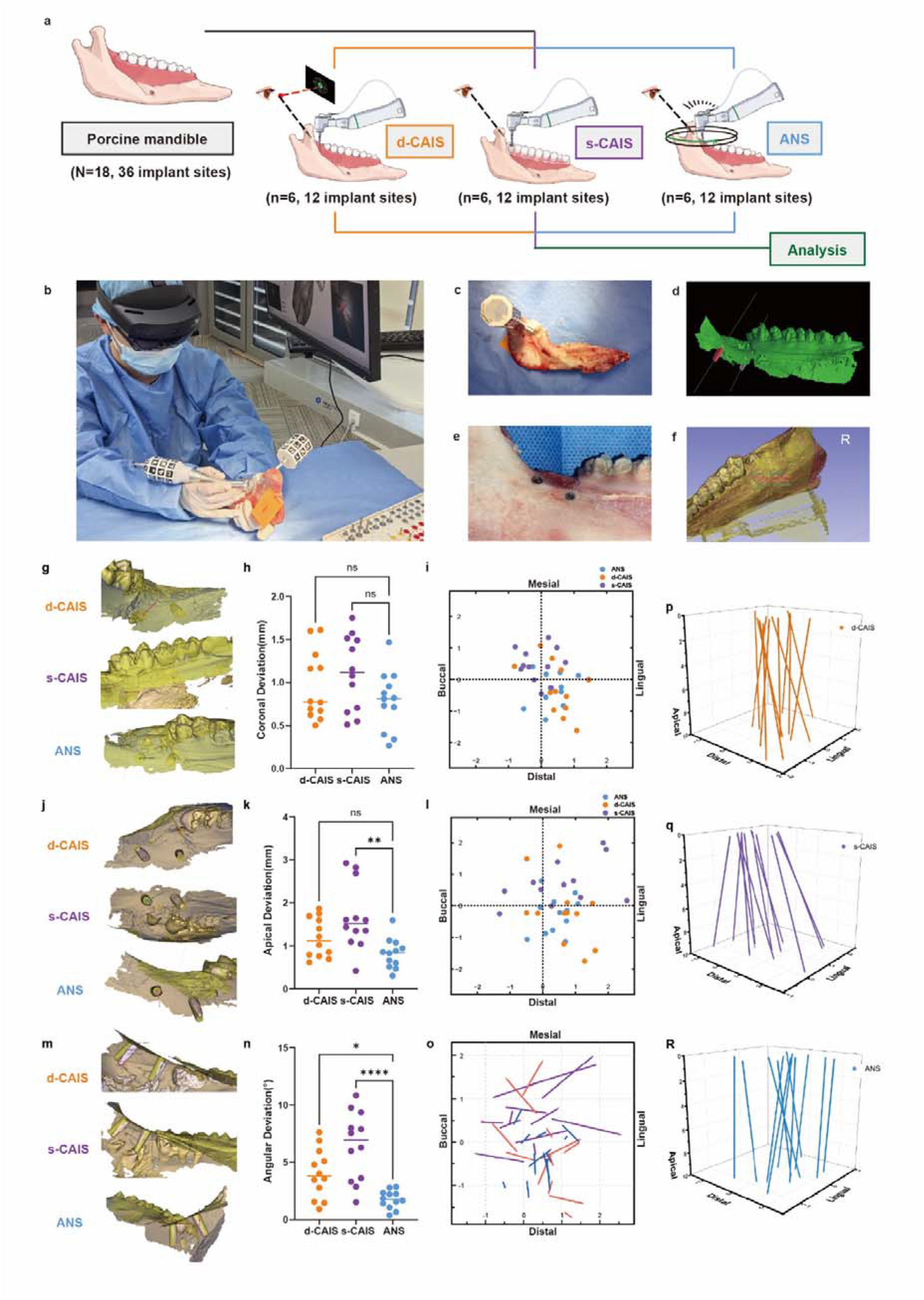
Accuracy evaluation of the augmented reality-based navigation system in a porcine mandible model. (a) Schematic illustration of the experimental design. Implant osteotomies were performed in porcine mandibles using dynamic computer-assisted implant surgery (d-CAIS), static computer-assisted implant surgery (s-CAIS), or the augmented reality-based navigation system (ANS), followed by postoperative accuracy analysis. (b) Representative intraoperative setup of ANS-guided drilling, in which the operator received real-time holographic guidance through an optical see-through head-mounted display. (c) Porcine mandible specimen with tracking markers attached for navigation and registration. (d) Three-dimensional reconstruction and virtual trajectory planning of the porcine mandible. (e) Representative drilled osteotomy sites in the porcine mandible. (f) Postoperative three-dimensional registration of the planned and actual drilling trajectories for accuracy assessment. (g) Representative reconstructed images showing coronal deviation in the d-CAIS, s-CAIS, and ANS groups. (h) Quantitative comparison of coronal deviation among the three groups. (i) Two-dimensional distribution map of coronal deviation in the mesial–distal and buccal–lingual directions. (j) Representative reconstructed images showing apical deviation in the d-CAIS, s-CAIS, and ANS groups. (k) Quantitative comparison of apical deviation among the three groups. (l) Two-dimensional distribution map of apical deviation in the mesial–distal and buccal–lingual directions. (m) Representative reconstructed images showing angular deviation in the d-CAIS, s-CAIS, and ANS groups. (n) Quantitative comparison of angular deviation among the three groups. (o) Directional distribution of angular deviation projected onto the mesial–distal and buccal–lingual plane. (p–r) Three-dimensional distribution of drilling trajectories in the d-CAIS, s-CAIS, and ANS groups, respectively. Data are presented as individual values with mean lines. Statistical significance was determined by one-way ANOVA followed by multiple-comparison testing. ns, not significant; *P < 0.05; **P < 0.01; ****P < 0.0001.

**Table 2.** Deviation statistics for indicator-assisted ANS, s-CAIS and d-CAIS in ex vivo porcine mandibles. Values summarise angular, three-dimensional entry-point and three-dimensional apical deviations;

| Group | ANS<br>(n=12) | s-CAIS<br>(n=12) | d-CAIS<br>(n=12) | P value (One-Way<br>ANOVA) |
| --- | --- | --- | --- | --- |
| **Angular deviation (degree)** |  |  |  |  |
| Mean±SD | 1.78±0.79 | 6.42±3.01 | 4.02±2.16 | **<0.001** |
| Median | 1.76 | 6.92 | 3.81 |  |
| Min-Max | 0.39-2.87 | 1.53-10.84 | 0.93-7.63 |  |
| **3D Entry point deviation (mm)** |  |  |  |  |
| Mean±SD | 0.79±0.34 | 1.11±0.44 | 0.96±0.40 | **0.157** |
| Median | 0.82 | 1.12 | 0.78 |  |
| Min-Max | 0.27-1.47 | 0.51-1.75 | 0.50-1.61 |  |
| **3D Apical deviation (mm)** |  |  |  |  |
| Mean±SD | 0.82±0.35 | 1.67±0.77 | 1.19±0.46 | **0.003** |
| Median | 0.84 | 1.52 | 1.12 |  |
| Min-Max | 0.31-1.60 | 0.42-2.93 | 0.62-1.87 |  |

One Way ANOVA was performed at $P$ value<0.05; ANS, augmented reality-based navigation system; s-CAIS, static Computer-Assisted Implant Surgery; d-CAIS, dynamic Computer-Assisted Implant Surgery; SD, standard deviation; Min, minimum value; Max, maximum value.

### ANS reduces perceived workload while maintaining placement accuracy

We next compared ANS with d-CAIS in postgraduate dental trainees. Placement accuracy was similar between systems, with no significant differences in coronal, apical or angular deviation. Mean deviations were consistently lower with ANS, but these differences did not reach statistical significance.

Questionnaire-based assessments indicated usability advantages for ANS. The SUS score was higher with ANS than with d-CAIS (73.13 vs. 68.88). NASA-TLX workload was lower with ANS (29.18 ± 11.44) than with d-CAIS (33.15 ± 13.67), indicating reduced perceived cognitive load (**Figure 4e-h**, **Table 3**). The differences in usability and workload were statistically significant (P = 0.014 and P = 0.031, respectively).

**Figure 4.**
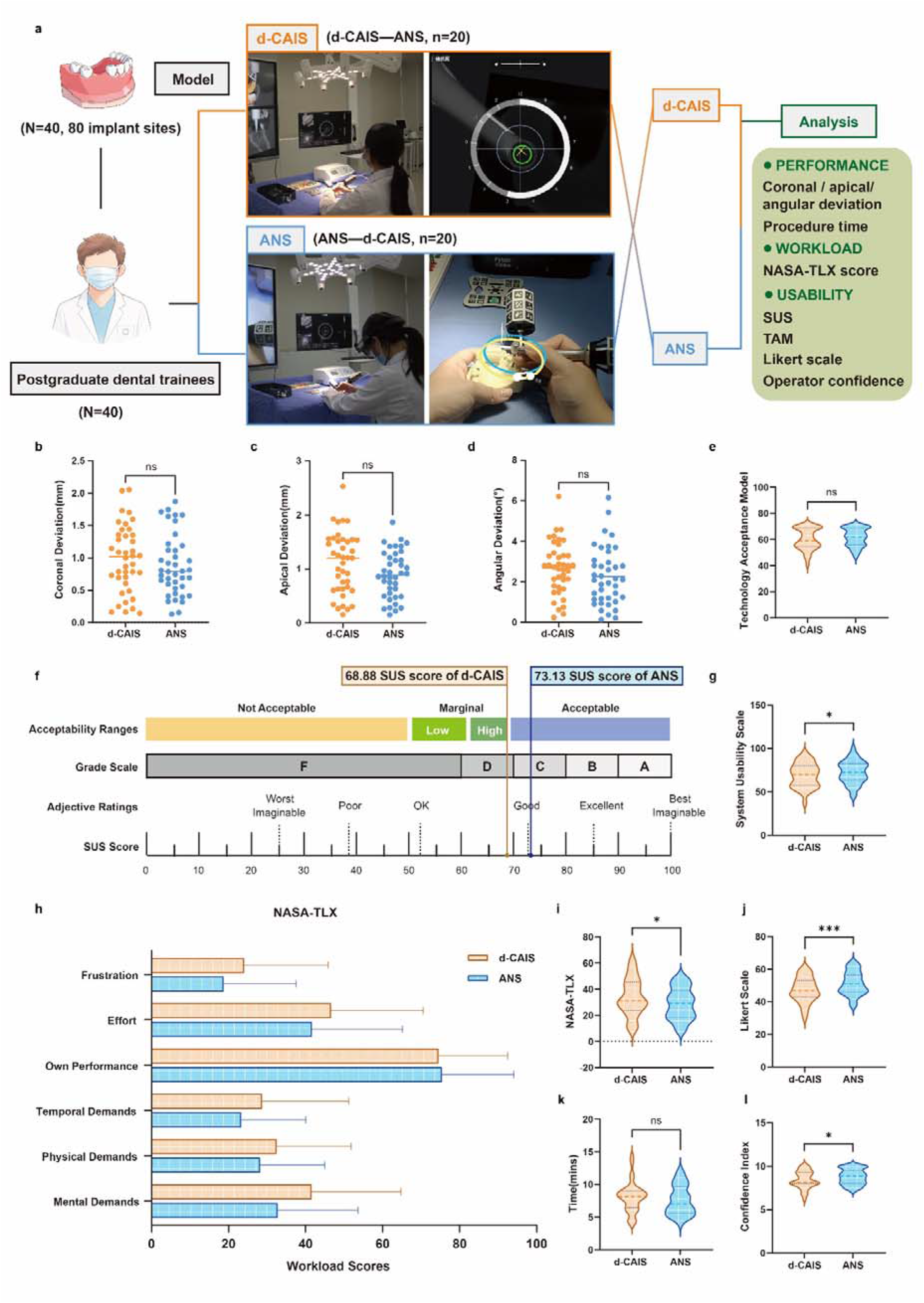
Accuracy, usability and workload comparison between ANS and d-CAIS in postgraduate dental trainees. (a) Schematic illustration of the crossover experimental design. The same cohort of postgraduate dental trainees performed implant osteotomy tasks using both dynamic computer-assisted implant surgery (d-CAIS) and the augmented reality-based navigation system (ANS). After completing each system-specific task, participants immediately evaluated the corresponding system before proceeding to the other navigation modality. Representative operative views and navigation interfaces are shown. (b–d) Comparison of coronal deviation, apical deviation, and angular deviation between d-CAIS and ANS. (e) Technology acceptance scores for the two systems. (f, g) System Usability Scale (SUS) benchmark and quantitative comparison of SUS scores. (h, i) NASA Task Load Index (NASA-TLX) subscale scores and overall workload scores. (j–l) Comparison of Likert-scale user experience, task completion time, and operator confidence index. Data are presented as individual values, violin plots, or mean ± SD as appropriate. Statistical significance was determined using paired t-tests or Wilcoxon matched-pairs signed-rank tests according to data distribution. ns, not significant; *P < 0.05; ***P < 0.001

**Table 3.**
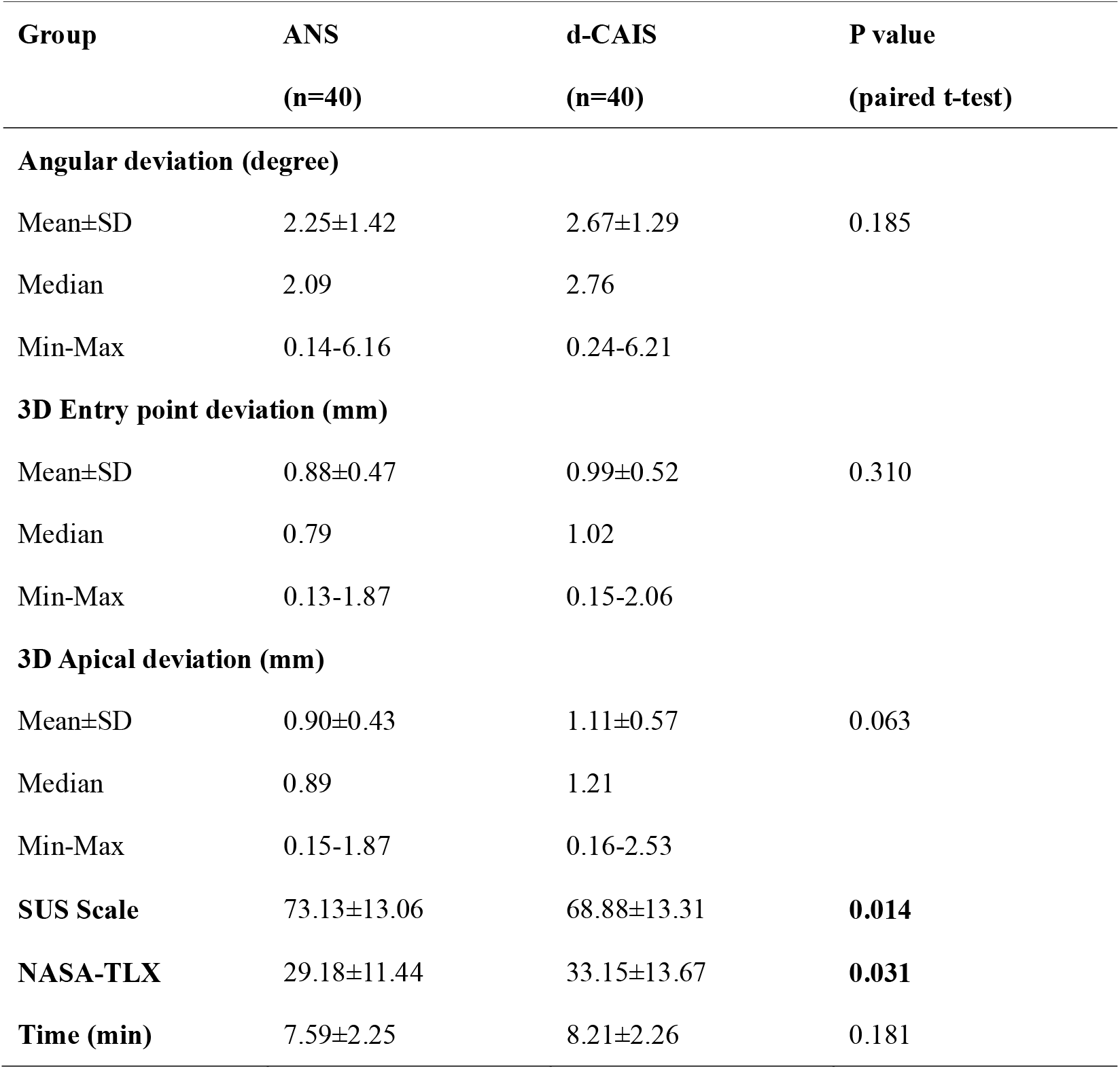
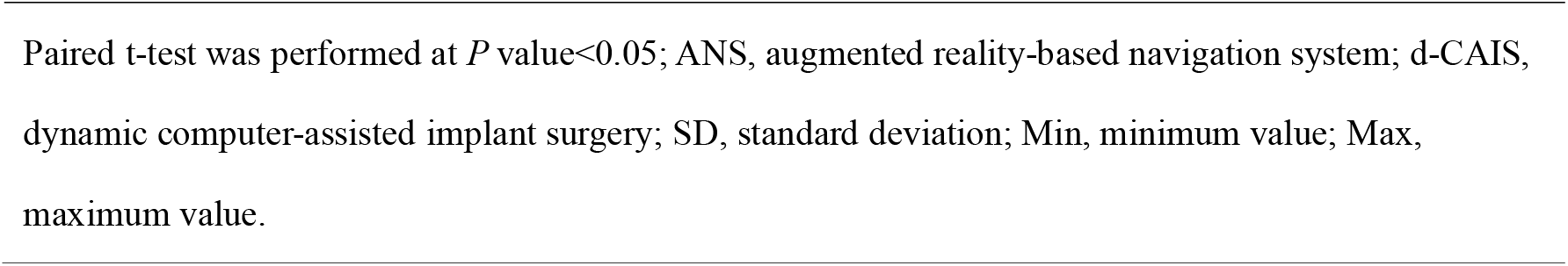
Deviation, usability, workload and time statistics for indicator-assisted ANS and d-CAIS in the postgraduate dental trainee crossover study. Values include placement deviations, SUS score, NASA-TLX score and procedure time; p values were calculated as indicated in the table.

| Group | ANS<br>(n=40) | d-CAIS<br>(n=40) | P value<br>(paired t-test) |
| --- | --- | --- | --- |
| **Angular deviation (degree)** |  |  |  |
| Mean±SD | 2.25±1.42 | 2.67±1.29 | 0.185 |
| Median | 2.09 | 2.76 |  |
| Min-Max | 0.14-6.16 | 0.24-6.21 |  |
| **3D Entry point deviation (mm)** |  |  |  |
| Mean±SD | 0.88±0.47 | 0.99±0.52 | 0.310 |
| Median | 0.79 | 1.02 |  |
| Min-Max | 0.13-1.87 | 0.15-2.06 |  |
| **3D Apical deviation (mm)** |  |  |  |
| Mean±SD | 0.90±0.43 | 1.11±0.57 | 0.063 |
| Median | 0.89 | 1.21 |  |
| Min-Max | 0.15-1.87 | 0.16-2.53 |  |
| **SUS Scale** | 73.13±13.06 | 68.88±13.31 | **0.014** |
| **NASA-TLX** | 29.18±11.44 | 33.15±13.67 | **0.031** |
| **Time (min)** | 7.59±2.25 | 8.21±2.26 | 0.181 |

Paired t-test was performed at $P$ value $<0.05$ ; ANS, augmented reality-based navigation system; d-CAIS, dynamic computer-assisted implant surgery; SD, standard deviation; Min, minimum value; Max, maximum value.

The Likert-scale assessment covered key components of navigation performance. Most operational indicators, including entry-point positioning, trajectory completion and interface complexity, did not differ significantly between ANS and d-CAIS. However, ANS was associated with better maintenance of visual focus on the surgical field (3.55 ± 1.47 vs. 2.28 ± 1.32, P < 0.001), lower perceived learning curve (3.15 ± 1.12 vs. 3.60 ± 1.22, *P = 0.020*), and less perceived interference with the drilling process (1.83 vs. 2.08, *P = 0.048*). Complete item-level responses are provided in **Table 4**. Operational confidence and real-time procedural adjustment showed favourable trends with ANS.

**Table 4.** Likert-scale assessment of navigation experience for ANS and d-CAIS. Scores are based on a 5-point Likert scale, with higher values indicating stronger agreement with each statement.

| No. | Likert Question | ANS<br>(n□=□40) | d-CAIS<br>(n□=□40) | Difference | *p* |
| --- | --- | --- | --- | --- | --- |
| 1 | It is easy to locate the starting point when using the navigation system. | 3.78 | 3.73 | 0.05 | 0.815 |
| 2 | It is convenient to confirm the drilling direction when using the system. | 4.25 | 3.98 | 0.28 | 0.162 |
| 3 | The system allows me to precisely adjust hand movements in real time. | 4.28 | 4.03 | 0.25 | 0.185 |
| 4 | During operation, my visual focus remains consistently on the surgical field. | 3.55 | 2.28 | 1.28 | **<0.001** |
| 5 | I can accurately complete the drilling trajectory with this system. | 4.28 | 4.03 | 0.25 | 0.058 |
| 6 | The navigation system interfered with my drilling process. | 1.83 | 2.08 | -0.25 | **0.048** |
| 7 | The guidance result after completion matches my drilling expectation. | 4.35 | 4.25 | 0.10 | 0.352 |
| 8 | I feel more confident when drilling using this system. | 4.40 | 4.20 | 0.20 | 0.058 |
| 9 | The system functions are intuitive and easy to understand. | 4.40 | 4.33 | 0.08 | 0.474 |
| 10 | The system interface is overly complex. | 2.13 | 2.18 | -0.05 | 0.720 |
| 11 | The navigation system is easy to operate. | 4.35 | 4.15 | 0.20 | 0.058 |

| No. | Likert Question | ANS<br>(n = 40) | d-CAIS<br>(n = 40) | Difference | *p* |
| --- | --- | --- | --- | --- | --- |
| 12 | I still need to learn many things before I can use the system proficiently. | 3.15 | 3.60 | -0.45 | **0.020** |
| 13 | Technical assistance is required when using this navigation system. | 3.43 | 3.53 | -0.10 | 0.533 |
| 14 | The various functions of the navigation system are well integrated. | 4.20 | 4.03 | 0.18 | 0.181 |

Values represent mean scores based on a 5-point Likert scale (1 = strongly disagree, 5 = strongly agree).
Paired t-test was performed at *P* value < 0.05; Bold *p* values indicate statistical significance (*p* < 0.05).

Participants also showed a higher confidence index when using ANS. Procedure time did not differ significantly between groups, although mean and median times were lower with ANS.

### ANS demonstrates preliminary clinical feasibility in mandibular first-molar placement

In the clinical mandibular first-molar case series, ANS achieved a mean angular deviation of 1.89 ± 1.64 degrees, a mean three-dimensional entry-point deviation of 0.65 ± 0.21 mm and a mean three-dimensional apical deviation of 0.90 ± 0.62 mm (**Figure 5i-n**, **Table 5**). The case distribution was balanced between left and right mandibular first molars, with four implants at site #36 and four at site #46. These results suggest that co-located AR guidance can be translated from model and ex vivo settings to selected posterior single-implant cases.

**Figure 5.**
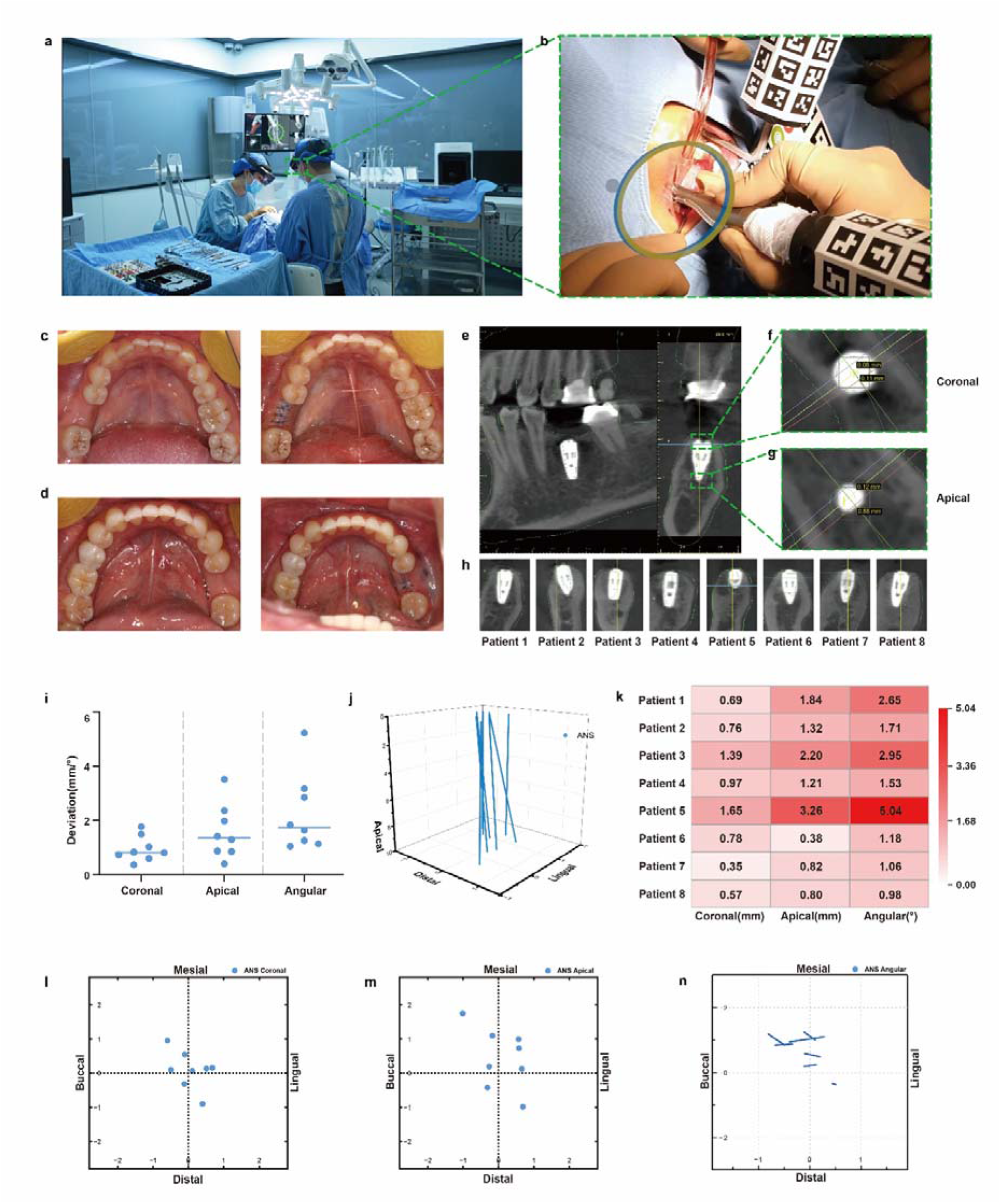
Preliminary clinical feasibility of ANS-guided implant surgery. (a) Operating-room setup during ANS-guided implant surgery. (b) Representative intraoperative view showing real-time holographic guidance during implant osteotomy. (c) Representative intraoral views before implant placement. (d) Representative intraoral views after ANS-guided implant placement. (e) Registration of the planned implant trajectory and the final postoperative CBCT, shown in representative coronal and sagittal views. (f, g) Transverse sections at the coronal and apical levels of the implant after registration, respectively. (h) Sagittal CBCT views of all eight clinical cases after ANS-guided implant placement. (i) Quantitative summary of coronal, apical, and angular deviations in the clinical cases. (j) Three-dimensional distribution of planned and placed implant trajectories. (k) Case-by-case heatmap showing coronal, apical, and angular deviations for each patient. (l) Directional distribution map of coronal deviation in the mesial–distal and buccal–lingual directions. (m) Directional distribution map of apical deviation in the mesial–distal and buccal–lingual directions. (n) Directional distribution map of angular deviation in the mesial–distal and buccal–lingual directions. Data are presented as individual values with mean lines.

**Table 5.** Clinical deviation statistics for ANS-guided mandibular first-molar implant placement. Values summarise angular, three-dimensional entry-point and three-dimensional apical deviations across eight single-implant cases.

| Group | ANS<br>(n=8) |
| --- | --- |
| **Angular deviation (degree)** |  |
| Mean±SD | 1.89±1.64 |
| Median | 1.45 |
| Min-Max | 0.50-5.04 |
| **3D Entry point deviation (mm)** |  |
| Mean±SD | 0.65±0.21 |
| Median | 0.59 |
| Min-Max | 0.38-1.09 |
| **3D Apical deviation (mm)** |  |
| Mean±SD | 0.90±0.62 |
| Median | 0.82 |
| Min-Max | 0.35-2.20 |
| ANS, augmented reality-based navigation system; SD, standard deviation; Min, minimum value;<br>Max, maximum value. |  |

## Discussion

This study developed and validated a holographic indicator-based AR navigation system for hard-tissue puncture surgery. In typodont models, ANS improved implant-placement angular and apical deviation compared with FHS. In ex vivo porcine mandibles, ANS showed higher angular and apical accuracy than s-CAIS and d-CAIS. In postgraduate dental trainees with no previous implant-placement experience, ANS achieved accuracy comparable to d-CAIS while reducing perceived workload and yielding higher self-reported visual-focus ratings. Finally, a preliminary mandibular first-molar case series showed clinically acceptable accuracy at matched posterior sites. Together, these findings support ANS as a practical action-guidance framework for AR surgical navigation.

The main distinction between ANS and many earlier AR-CAS systems is the role assigned to visual information. Traditional systems largely augment information by displaying trajectories, coordinates or anatomical relationships on monitors or virtual panels 34-37. The surgeon must then integrate these data with the surgical field. ANS instead converts navigation error into co-located holographic action cues. This design may shorten the cognitive pathway from perception to execution, which is consistent with the lower NASA-TLX scores and higher self-reported visual-focus ratings observed in this study. Compared with published ex vivo AR-CAS studies, ANS also achieved favourable angular accuracy (Table 6). Although no universally accepted thresholds define clinically significant angular deviation in implant surgery, angular error is clinically relevant because it contributes to apical displacement and may compromise anatomical safety margins. For a 10-mm implant, angular deviations of 3°, 5°, and 6° correspond approximately to lateral apical displacements of 0.52 mm, 0.87 mm, and 1.05 mm, respectively. Therefore, the reduction in angular deviation observed with ANS in the ex vivo experiment, particularly compared with s-CAIS, may be clinically meaningful in anatomically constrained sites. In contrast, the smaller angular improvement observed in typodont models, although statistically significant, may have more limited direct clinical impact. The clinical case series demonstrated deviations within commonly reported ranges for guided implant surgery, supporting feasibility, but controlled clinical studies are required to determine comparative clinical significance.

**Table 6.** Accuracy comparison with published ex vivo AR-CAS studies. Coronal, apical and angular deviations are reported as published by each study; differences in models, surgical sites, registration methods and measurement protocols limit direct statistical comparison.

| Research | Coronal (mm) | Apical (mm) | Angular (°) |
| --- | --- | --- | --- |
| Lin et al. <sup>38</sup> | 0.46 ± 0.20 | 1.23 ± 0.42 | 3.33 ± 1.42 |
| Katić et al. <sup>39</sup> | 1.10 – 2.48 | 1.10 – 2.48 | ≈2 |
| Jiang et al. <sup>40</sup> | 1.25 – 1.68 | 1.34 – 1.75 | 3.37 – 5.04 |
| Kivovics et al.<sup>41</sup> | 1.27 ± 0.40 | 1.34 ± 0.41 | 4.09 ± 2.79 |
| Ma et al.<sup>42</sup> | 1.25 ± 0.33 | 1.25 ± 0.33 | 4.03 ± 1.56 |
| Lin et al.<sup>43</sup> | 0.69 ± 0.25 | 0.79 ± 0.23 | 1.85 ± 0.61 |
| Fan et al.<sup>44</sup> | 1.51 ± 0.16 | 1.54 ± 0.14 | 3.47 ± 0.34 |
| **Our** | **0.79±0.34** | **0.82±0.35** | **1.78±0.80** |

The clinical first-molar findings should be interpreted as preliminary evidence of feasibility rather than comparative efficacy. This distinction is important because the present clinical series did not include contemporaneous s-CAIS or d-CAIS controls. Recent meta-analyses indicate that static and dynamic CAIS both improve transfer accuracy relative to freehand placement, with no clinically relevant difference between static and dynamic approaches in clinical comparative studies^45^. Reported pooled or weighted estimates for guided implant placement commonly fall around 1.1-1.2 mm at the coronal platform, 1.0-1.4 mm at the apex and 3.5-4.1 degrees for angular deviation^45,46^. A posterior single-tooth d-CAIS clinical series, which is more anatomically comparable to the present first-molar cases than mixed-site studies, reported platform, apical and angular deviations of 0.90 ± 0.46 mm, 1.04 ± 0.47 mm and 2.54 ± 1.21 degrees, respectively^47^. Within this context, the present #36/#46 ANS cases showed comparable or lower mean platform and apical deviations, but the small sample size and indirect nature of these comparisons preclude claims of superiority.

Our findings extend this earlier randomized evidence by examining a different interface paradigm in which navigation discrepancies are translated into co-located action cues rather than primarily visualized as spatial information ^20^. This shift from information display to action guidance may have practical implications for hard-tissue surgery. By reducing reliance on external screens, co-located indicators may improve posture, preserve attention within the operative field and simplify training for procedures that require three-dimensional spatial control. These advantages may be especially relevant in posterior implant placement, where access, visibility and angulation control can be difficult. However, this potential should be confirmed in procedure-specific clinical studies before broad implementation.

The distributed visible-light architecture avoided reliance on infrared tracking and was designed to reduce potential optical interference with the AR display. The unified visual-recognition framework coordinated the patient, instrument and display coordinate systems within a shared visual space, enabling stable spatial anchoring of the indicators. Edge computing supported low-latency tracking and rendering, which are necessary for real-time guidance.

Several limitations remain. First, the clinical case series was small, non-randomised and restricted to mandibular first-molar sites with sufficient bone volume. It therefore cannot establish comparative effectiveness against s-CAIS or d-CAIS. Second, generalisability of the registration requires further testing. Marker-based registration performed well in dental models and selected clinical cases, but its robustness in regions with thick soft-tissue coverage or complex bone morphology, such as the pelvis or scapula, remains uncertain ^48^. Marker-free registration based on anatomical surface contours or preplaced fiducials may be needed. Third, prolonged OST-HMD use may cause visual fatigue or spatial-perception errors ^49,50^. Device weight, field-of-view limits and residual vergence-accommodation conflict should be addressed through hardware and software optimisation. Finally, integration of ANS into routine clinical workflow has not yet been validated in larger operating-room cohorts.

Taken together, these findings support a co-located action-guidance framework in which AR navigation moves beyond information display toward directly actionable intraoperative guidance. In dental implant placement, ANS demonstrated improved accuracy over freehand placement, favourable performance relative to existing navigation approaches in ex vivo testing, reduced perceived workload in trainees, and preliminary clinical feasibility. The framework may be adaptable to other hard-tissue procedures that require stringent spatial control, including selected spinal, orthopaedic and neurosurgical applications. Its broader value will depend on registration robustness, device ergonomics and validation in larger, controlled clinical workflows.

With these boundaries, ANS suggests a direction for surgical navigation in which AR systems move from presenting complex information toward delivering intuitive, action-level guidance. Further optimisation and prospective clinical validation are needed before this approach can be translated into routine surgical practice.

## Methods

### Study design and ANS implementation

#### Study design

This multistage translational study comprised randomized typodont and ex vivo porcine experiments, a randomized crossover study in postgraduate dental trainees, and an uncontrolled clinical case series. The study was conducted from July 2024 to March 2026 at West China Hospital of Stomatology, Sichuan University, China.

#### ANS architecture and workflow

The ANS comprised a self-developed optical see-through head-mounted display (OST-HMD), a distributed vision-tracking system consisting of six visible-light cameras with overlapping fields of view, a cart-mounted GPU-accelerated computing workstation for real-time visual tracking and navigation processing, and ArUco-based visual trackers attached to the patient and surgical handpiece (**Figure 1e**).

The patient tracker was connected to the dentition through a dedicated fixation device and secured using a self-curing temporization material (Protemp™ 4 Temporization Material; 3M ESPE, Seefeld, Germany). The handpiece tracker was rigidly connected to the surgical handpiece (Ti-Max X-DSG20L; NSK, Japan). Image acquisition, visual-marker recognition, pose estimation and real-time tracking were performed by the cart-mounted computing workstation.

A web-based planning module supported CBCT reconstruction and implant planning. A C++ navigation module estimated the poses of the patient and surgical instrument, established spatial correspondence among the patient, surgical instrument, OST-HMD and virtual surgical plan, and continuously calculated lateral displacement, angular deviation, remaining drilling depth and safety margins. A Unity/C# visualization module rendered the co-located lateral, angular and depth indicators through the OST-HMD. Interactive CBCT reslicing allowed the surgeon to reposition the sectioning plane and inspect the anatomy from different orientations.

#### Spatial registration and instrument calibration

Before navigation, the six-camera system detected ArUco markers mounted on the OST-HMD, patient tracker and handpiece tracker. Recognition of these markers established a common spatial reference among the camera system, head-mounted display, patient and surgical instrument.

Patient-to-image registration was performed using predefined dental cusp landmarks corresponding to the preoperative digital model. The physical cusp landmarks were identified intraoperatively and matched with their corresponding points in the virtual model to establish the spatial relationship between the patient and the preoperative plan.

Handpiece and tool calibration was performed through multi-view visual recognition by the camera system. The handpiece was presented within the camera field of view, allowing the system to recognize the tracker and corresponding instrument geometry from multiple orientations without conventional manual point-based calibration. When a drill was changed, the handpiece was repositioned within the calibration area for automatic recognition and updating of the corresponding tool geometry. After each drill change, patient registration was reverified using the predefined dental cusp landmarks before navigation resumed.

### Accuracy evaluation

#### Preliminary accuracy validation in typodont models

Thirty identical bilateral first-molar edentulous typodont models (60 implant sites; tolerance <0.1 mm) were randomly assigned to an AR-based navigation group (ANS, n=15 models, 30 sites) or a freehand surgery group (FHS, n=15 models, 30 sites). All models were scanned, and an average model was selected as the planning dataset. Implant positions were planned in the ANS software and used as the unified planning standard for both groups. One experienced surgeon performed all drilling procedures under ANS or FHS guidance, using the same method bilaterally within each model. Straumann implants (4.1 × 10 mm; Straumann, Switzerland) were placed according to the standard implantation workflow. Postoperative scan-body STL files were obtained for accuracy assessment (**Figure 2a-g**).

#### Preclinical accuracy validation in porcine mandibles

To evaluate ANS under clinically relevant ex vivo conditions, 18 fresh porcine mandibles from 7-to 9-month-old animals were obtained from a local market and randomly assigned to ANS, s-CAIS, or d-CAIS (6 mandibles per group). All mandibles underwent CBCT scanning with a 3D Accuitomo system (Morita; 85 kV, 6 mA, 17.5-second exposure; 170 × 120-mm field of view), and intraoral scans were also obtained for surface registration.

Implant sites were planned along the external oblique line adjacent to the mandibular ramus and perpendicular to the bone surface by an oral-imaging operator who did not participate in surgery.

All procedures were performed by the same implant specialist with four years of clinical experience. The surgeon was experienced in freehand and static CAIS procedures and completed multiple model surgeries with ANS. Interventions were performed in random order over two days to reduce fatigue and sequence effects. All drilling was performed at 800 rpm with external water cooling. Postoperative datasets were obtained for accuracy assessment (**Figure 3a-g**).

#### Usability evaluation

A randomized crossover study was conducted among postgraduate dental trainees to compare ANS with conventional dynamic navigation in terms of placement accuracy, usability, cognitive workload, and navigation experience. Forty postgraduate dental trainees were recruited to evaluate system practicality using a bilateral mandibular first-molar edentulous typodont model. All participants had at least two years of clinical experience but had not independently completed implant placement. Ethical approval for this study was obtained from the Ethics Committee of West China Hospital of Stomatology, Sichuan University, and written informed consent was obtained from all participants. Participants were randomly assigned to two sequence groups. One group completed ANS-guided placement, completed the questionnaires, rested for 10 min and then completed d-CAIS-guided placement. The other group completed the same procedures in the reverse order. To minimize recall and order effects, the sequence of system use was randomized among participants. Each participant placed two implants with the starting side randomly assigned.

All training models were scanned preoperatively, with measured tolerances of less than 0.01 mm. An average model was selected for ANS planning, and cusp points were marked for registration.

All participants completed both navigation workflows during the same afternoon. In total, 80 implants (Straumann, 4.1 × 10 mm; Straumann, Switzerland) were placed, 80 questionnaires were collected and procedure time was recorded for each placement. After implantation, scan-body reconstruction was performed and STL files were exported.

#### Clinical feasibility assessment

To preliminarily evaluate the clinical feasibility and placement accuracy of ANS in a clinical setting, we conducted an uncontrolled case series of eight patients requiring single mandibular first-molar implant placement. Four implants were placed at site #36 and four at site #46. Patient characteristics and implant specifications are provided in **Table 7**. The study was approved by the Ethics Committee of West China Hospital of Stomatology, Sichuan University (No. WCHS-CRSE-2024-177-R1-P), and written informed consent was obtained from all participants. The clinical study was registered and filed in the Medical Research Registration and Filing Information System (filing no. MR-51-25-065783). Eligible patients were adults with good general and oral health, sufficient bone volume for implant placement and no requirement for additional hard- or soft-tissue augmentation.

**Table 7.**
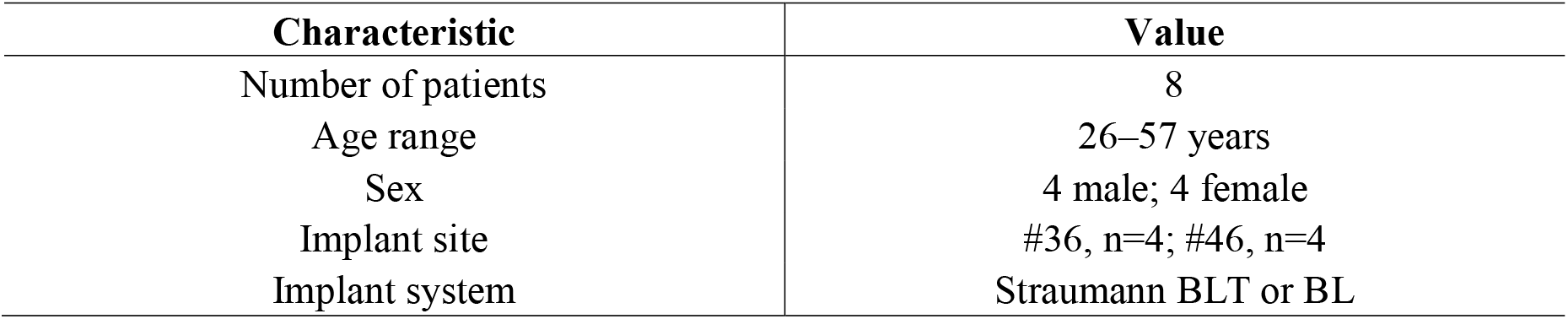
Baseline characteristics of participants in the clinical feasibility study.

All patients underwent preoperative CBCT scanning. Virtual implant planning was performed in the ANS software according to patient-specific anatomy and prosthetically driven requirements. Implant dimensions were selected on the basis of available bone volume, and a safety margin was incorporated to evaluate the relationship between the planned implant and adjacent anatomical structures.

During surgery, tracking markers were fixed intraorally and to the surgical handpiece. After automatic recognition by the camera system, spatial registration between the patient, handpiece and virtual plan was established and verified using predefined validation points. Osteotomy preparation and implant placement were then performed under continuous AR guidance. The holographic indicator provided real-time feedback on drilling position, depth and angulation directly within the operative field. Postoperative CBCT was obtained for three-dimensional comparison with the planned implant position (**Figure 5a-h**).

#### Outcome measures and statistical analysis

The primary accuracy outcomes were coronal, apical, and angular deviations between the planned and actual implant trajectories. Coronal and apical deviations were defined as the three-dimensional distances between the corresponding reference points, and angular deviation as the angle between the trajectory axes (**Figure 2b**). For the typodont and porcine experiments, planned and postoperative datasets were rigidly registered in 3D Slicer using stable anatomical structures. Clinical datasets were registered and measured in Blue Sky Plan. Secondary outcomes in the crossover study included procedure time, technology acceptance, System Usability Scale scores, NASA Task Load Index scores, navigation-experience ratings, and operator confidence.

Statistical analyses were performed using SAS. Normality was assessed using the Shapiro–Wilk test. Independent typodont groups were compared using unpaired tests, and the three porcine groups using one-way analysis of variance with post hoc comparisons. Within-participant outcomes in the crossover study were analysed using paired tests. Clinical outcomes were summarised descriptively. All tests were 2-sided, with P < 0.05 considered statistically significant.

## Author contributions

TT and XC supervised and reviewed the study. TT also provided the study materials, data, computing resources, and secured financial support for the project. DZ contributed to data curation, formal analysis, investigation, project administration, methodology, software development, validation, visualisation, and drafting of the manuscript. YQ was responsible for formal analysis, methodology, software development, supervision, and manuscript review and editing. YL contributed to data curation. YG, XRL and XTL contributed to resources and validation. DZ, YQ, and YL accessed and verified the raw data. HC contributed to supervision, and manuscript review and editing. All authors reviewed the manuscript and had final responsibility for the decision to submit for publication.

## Data availability

De-identified data supporting the findings of this study are available in Mendeley Data http://doi.org/10.17632/x6p8k647sz.151. Additional clinical data that could compromise participant confidentiality are available from the corresponding author upon reasonable request and subject to institutional and ethical approval.

## Code availability

Relevant code or implementation details required to evaluate the findings of this study will be made available to editors and reviewers upon request. Following publication, access by qualified researchers will be subject to applicable institutional and intellectual property requirements.

## Acknowledgements

This study was supported by the National Natural Science Foundation of China (82201027), the Young Elite Scientists Sponsorship Program by CAST (YESS20240133), and the Sichuan Science and Technology Program (2024NSFSC1591). The funders had no role in the study design; data collection, analysis, or interpretation; preparation of the manuscript; or the decision to submit the manuscript for publication.

## References

1 Clement, H., Heidari, N., Grechenig, W., Weinberg, A. M. & Pichler, W. Drilling, not a benign procedure: laboratory simulation of true drilling depth. Injury 43, 950–952 (2012). 10.1016/j.injury.2011.11.017

2 Klok, J. W., et al. Design and In Vitro Validation of an Orthopaedic Drill Guide for Femoral Stem Revision in Total Hip Arthroplasty. IEEE J Transl Eng Health Med 12, 340–347 (2024). 10.1109/jtehm.2024.3365300

3 Neal, K. M., Gallaher, H. M., Thompson, A. & Kerby, M. D. The effect of an aiming device on the accuracy of humeral transcondylar screw placement. Vet Surg 52, 538–544 (2023). 10.1111/vsu.13952

4 Abood, A., et al. Autofluorescence-Guided Total Thyroidectomy in Low-Volume, Nonparathyroid Institutions. JAMA Netw Open 7, e2411384 (2024). 10.1001/jamanetworkopen.2024.11384

5 van Keulen, S., et al. The Clinical Application of Fluorescence-Guided Surgery in Head and Neck Cancer. J Nucl Med 60, 758–763 (2019). 10.2967/jnumed.118.222810

6 Guenette, J. P., Tuncali, K., Himes, N., Shyn, P. B. & Lee, T. C. Percutaneous Image-Guided Cryoablation of Head and Neck Tumors for Local Control, Preservation of Functional Status, and Pain Relief. AJR Am J Roentgenol 208, 453–458 (2017). 10.2214/AJR.16.16446

7 Guha, D. et al. Spinal intraoperative three-dimensional navigation: correlation between clinical and absolute engineering accuracy. Spine J 17, 489–498 (2017). 10.1016/j.spinee.2016.10.020

8 Gueziri, H.-E., Georgiopoulos, M., Santaguida, C. & Collins, D. L. Ultrasound-based navigated pedicle screw insertion without intraoperative radiation: feasibility study on porcine cadavers. Spine J 22, 1408–1417 (2022). 10.1016/j.spinee.2022.04.014

9 Mandel, W., Oulbacha, R., Roy-Beaudry, M., Parent, S. & Kadoury, S. Image-Guided Tethering Spine Surgery With Outcome Prediction Using Spatio-Temporal Dynamic Networks. IEEE Trans Med Imaging 40, 491–502 (2021). 10.1109/TMI.2020.3030741

10 Lee, J. Y. K., et al. Near-infrared fluorescent image-guided surgery for intracranial meningioma. J Neurosurg 128, 380–390 (2018). 10.3171/2016.10.JNS161636

11 Mundinano, I.-C., Flecknell, P. A. & Bourne, J. A. MRI-guided stereotaxic brain surgery in the infant and adult common marmoset. Nat Protoc 11, 1299–1308 (2016). 10.1038/nprot.2016.076

12 Dixon, B. J. et al. Surgeons blinded by enhanced navigation: the effect of augmented reality on attention. Surg Endosc 27, 454–461 (2013). 10.1007/s00464-012-2457-3

13 Waelkens, P., van Oosterom, M. N., van den Berg, N. S., Navab, N. & van Leeuwen, F. W. B. in Radioguided Surgery: Current Applications and Innovative Directions in Clinical Practice (eds Ken Herrmann, Omgo E. Nieweg, & Stephen P. Povoski) 57–73 (Springer International Publishing, 2016).

14 Frisk, H. et al. Feasibility and Accuracy of Thoracolumbar Pedicle Screw Placement Using an Augmented Reality Head Mounted Device. Sensors 22, 522 (2022).

15 Müller, F. et al. Augmented reality navigation for spinal pedicle screw instrumentation using intraoperative 3D imaging. The Spine Journal 20, 621–628 (2020). 10.1016/j.spinee.2019.10.012

16 Tu, P., Wang, H., Joskowicz, L. & Chen, X. A multi-view interactive virtual-physical registration method for mixed reality based surgical navigation in pelvic and acetabular fracture fixation. International journal of computer assisted radiology and surgery 18, 1715–1724 (2023). 10.1007/s11548-023-02884-4

17 Tu, P. et al. Augmented reality based navigation for distal interlocking of intramedullary nails utilizing Microsoft HoloLens 2. Computers in biology and medicine 133, 104402 (2021). 10.1016/j.compbiomed.2021.104402

18 Kriechling, P., et al. Augmented reality through head-mounted display for navigation of baseplate component placement in reverse total shoulder arthroplasty: a cadaveric study. Archives of Orthopaedic and Trauma Surgery 143, 169–175 (2023). 10.1007/s00402-021-04025-5

19 Burström, G., Nachabe, R., Persson, O., Edström, E. & Elmi Terander, A. Augmented and Virtual Reality Instrument Tracking for Minimally Invasive Spine Surgery: A Feasibility and Accuracy Study. Spine (Phila Pa 1976) 44, 1097–1104 (2019). 10.1097/brs.0000000000003006

20 Li, Y., et al. Accuracy and efficiency of drilling trajectories with augmented reality versus conventional navigation randomized crossover trial. npj Digital Medicine 7, 316 (2024). 10.1038/s41746-024-01314-2

21 Pellegrino, G. et al. Augmented reality for dental implantology: a pilot clinical report of two cases. BMC oral health 19, 158 (2019). 10.1186/s12903-019-0853-y

22 Wei, B., et al. Augmented reality in preoperative anterolateral thigh flap perforators positioning: A pilot diagnostic study. Oral Oncology 162, 107189 (2025). 10.1016/j.oraloncology.2025.107189

23 Liu, L., et al. A mixed reality-based navigation method for dental implant navigation method: A pilot study. Computers in biology and medicine 154, 106568 (2023). 10.1016/j.compbiomed.2023.106568

24 Engelschalk, M., Al Hamad, K. Q., Mangano, R., Smeets, R. & Molnar, T. F. Dental implant placement with immersive technologies: A preliminary clinical report of augmented and mixed reality applications. The Journal of prosthetic dentistry 133, 346–351 (2025). 10.1016/j.prosdent.2024.02.017

25 Hong, W., et al. A Self-Developed Mobility Augmented Reality System Versus Conventional X-rays for Spine Positioning in Intraspinal Tumor Surgery: A Case-Control Study. Neurospine 21, 984–993 (2024). 10.14245/ns.2448188.094

26 Kivovics, M., Takács, A., Pénzes, D., Németh, O. & Mijiritsky, E. Accuracy of dental implant placement using augmented reality-based navigation, static computer assisted implant surgery, and the free-hand method: An in vitro study. Journal of dentistry 119, 104070 (2022). 10.1016/j.jdent.2022.104070

27 Fan, X. et al. A novel mixed reality-guided dental implant placement navigation system based on virtual-actual registration. Computers in biology and medicine 166, 107560 (2023). 10.1016/j.compbiomed.2023.107560

28 Kramida, G. Resolving the Vergence-Accommodation Conflict in Head-Mounted Displays. IEEE transactions on visualization and computer graphics 22, 1912–1931 (2016). 10.1109/TVCG.2015.2473855

29 Wolf, J., et al. How different augmented reality visualizations for drilling affect trajectory deviation, visual attention, and user experience. International journal of computer assisted radiology and surgery 18, 1363–1371 (2023). 10.1007/s11548-022-02819-5

30 Jin, X. et al. in Proceedings of the 2025 Symposium on Eye Tracking Research and Applications Article 83 (Association for Computing Machinery, 2025).

31 Baumeister, J., et al. Cognitive Cost of Using Augmented Reality Displays. IEEE transactions on visualization and computer graphics 23, 2378–2388 (2017). 10.1109/tvcg.2017.2735098

32 Bapna, T., Valles, J., Leng, S., Pacilli, M. & Nataraja, R. M. Eye-tracking in surgery: a systematic review. ANZ Journal of Surgery 93, 2600–2608 (2023). 10.1111/ans.18686

33 Popov, V. et al. in *Proceedings of the 2024 CHI Conference on Human Factors in Computing Systems* Article 459 (Association for Computing Machinery, Honolulu, HI, USA, 2024).

34 Martin-Gomez, A., et al. STTAR: Surgical Tool Tracking Using Off-the-Shelf Augmented Reality Head-Mounted Displays. IEEE transactions on visualization and computer graphics 30, 3578–3593 (2024). 10.1109/tvcg.2023.3238309

35 Hu, X., Baena, F. R. Y. & Cutolo, F. Head-Mounted Augmented Reality Platform for Markerless Orthopaedic Navigation. IEEE J Biomed Health Inform 26, 910–921 (2022). 10.1109/jbhi.2021.3088442

36 Wang, E., Liu, Y., Tu, P., Taylor, Z. A. & Chen, X. Video-Based Soft Tissue Deformation Tracking for Laparoscopic Augmented Reality-Based Navigation in Kidney Surgery. IEEE Trans Med Imaging 43, 4161–4173 (2024). 10.1109/tmi.2024.3413537

37 von Atzigen, M., et al. Marker-free surgical navigation of rod bending using a stereo neural network and augmented reality in spinal fusion. Med Image Anal 77, 102365 (2022). 10.1016/j.media.2022.102365

38 Lin, Y. K., Yau, H. T., Wang, I. C., Zheng, C. & Chung, K. H. A novel dental implant guided surgery based on integration of surgical template and augmented reality. Clinical implant dentistry and related research 17, 543–553 (2015). 10.1111/cid.12119

39 Katić, D., et al. A system for context-aware intraoperative augmented reality in dental implant surgery. International journal of computer assisted radiology and surgery 10, 101–108 (2015). 10.1007/s11548-014-1005-0

40 Jiang, W., et al. Evaluation of the 3D Augmented Reality-Guided Intraoperative Positioning of Dental Implants in Edentulous Mandibular Models. The International journal of oral & maxillofacial implants 33, 1219–1228 (2018). 10.11607/jomi.6638

41 Kivovics, M., Takács, A., Pénzes, D., Németh, O. & Mijiritsky, E. Accuracy of dental implant placement using augmented reality-based navigation, static computer assisted implant surgery, and the free-hand method: An in vitro study. Journal of dentistry 119, 104070 (2022). 10.1016/j.jdent.2022.104070

42 Ma, L., et al. Augmented reality surgical navigation with accurate CBCT-patient registration for dental implant placement. Medical & biological engineering & computing 57, 47–57 (2019). 10.1007/s11517-018-1861-9

43 Lin, C., et al. Application of mixed reality-based surgical navigation system in craniomaxillofacial trauma bone reconstruction. Hua Xi Kou Qiang Yi Xue Za Zhi 40, 676–684 (2022). 10.7518/hxkq.2022.06.008

44 Fan, X. et al. A novel mixed reality-guided dental implant placement navigation system based on virtual-actual registration. Computers in biology and medicine 166, 107560 (2023). 10.1016/j.compbiomed.2023.107560

45 Reiff, F. S., Kroeger, A., Roehling, S., Reiff, C. & Kebschull, M. Accuracy of Freehand, Static, and Dynamic Computer-Assisted Implant Placement: *A Systematic Review and Meta-Analysis*. J Periodontal Res 61, 111–137 (2026). 10.1111/jre.70047

46 Schnutenhaus, S., Edelmann, C., Knipper, A. & Luthardt, R. G. Accuracy of Dynamic Computer-Assisted Implant Placement: A Systematic Review and Meta-Analysis of Clinical and In Vitro Studies. Journal of clinical medicine 10, 704 (2021).

47 Wu, B.-Z., Xue, F., Ma, Y. & Sun, F. Accuracy of automatic and manual dynamic navigation registration techniques for dental implant surgery in posterior sites missing a single tooth: A retrospective clinical analysis. Clinical oral implants research 34, 221–232 (2023). 10.1111/clr.14034

48 Carbone, M., et al. Architecture of a Hybrid Video/Optical See-through Head-Mounted Display-Based Augmented Reality Surgical Navigation Platform. Information 13, 81 (2022).

49 Condino, S., Carbone, M., Piazza, R., Ferrari, M. & Ferrari, V. Perceptual Limits of Optical See-Through Visors for Augmented Reality Guidance of Manual Tasks. IEEE Trans Biomed Eng 67, 411–419 (2020). 10.1109/tbme.2019.2914517

50 Birlo, M., Edwards, P. J. E., Clarkson, M. & Stoyanov, D. Utility of optical see-through head mounted displays in augmented reality-assisted surgery: A systematic review. Med Image Anal 77, 102361 (2022). 10.1016/j.media.2022.102361

51 Tian, T., Zhao, D. Accuracy Metrics and User-Experience Evaluations for an Augmented Reality Surgical Navigation (ANS), Mendeley Data, V1, (2025). 10.17632/x6p8k647sz.1.

